# Prospective Evaluation of AI Risk Stratification for Triaging Expedited Screening Mammogram Interpretation

**DOI:** 10.1101/2025.10.10.25337626

**Authors:** Maggie Chung, Eric Davis, Heather Greenwood, Jessica Hayward, Shinn-Huey Chou, Bonnie Joe, Loretta Strachowski, Tatiana Kelil, Rita Freimanis, Elissa Price, Kimberly Ray, Amie Lee, Adam Yala

## Abstract

**PURPOSE:** To prospectively evaluate the feasibility and performance of expedited screening mammogram interpretation for women identified as high-risk by a deep learning risk model.

**METHODS AND MATERIALS:** This HIPAA-compliant, IRB-approved prospective controlled study was conducted at an urban safety-net facility. The Mirai breast cancer risk model was retrospectively validated on 114,229 local mammograms (2006–2023) to identify the top 10% of 1-year breast cancer risk threshold. During the prospective study (12/2024-6/2025), Mirai 1-year risk scores were generated in real-time. On enrollment days, high-risk women were consented and offered immediate screening mammogram interpretation. Patients assessed as BI-RADS 0 were offered same-day diagnostic evaluation when feasible. Outcomes included feasibility of immediate interpretation, time to screening result (Ts), diagnostic evaluation (Td), and biopsy (Tb), as well as cancer detection rate (CDR). Comparisons were made with high-risk controls on non-enrollment days.

**RESULTS:** Among 4,145 screening mammograms, Mirai flagged 525 (12.7%) as high-risk; 973 (23.5%) were performed on enrollment days with 115 (11.8%) flagged as high-risk. Of 100 women who consented, 94% received immediate reads. Thirty-one were assessed as BI-RADS 0; 30 underwent diagnostic imaging (26 same-day). Thirteen biopsies yielded 6 malignant, 2 high-risk, and 5 benign lesions. The CDR in high-risk expedited women was 60/1,000 (95% CI, 22.3–126.0) compared with 2.3/1,000 (95% CI, 0.3–8.4) in non-high-risk women (odds ratio 27.1; p<0.001). Median Ts, Td, and Tb were significantly shorter in expedited patients vs. high-risk controls (13.0 min vs. 191.9 min; 1.3 hrs vs. 852.8 hrs; 20.1 vs. 59.0 days; all p<0.001). For screen-detected cancers, expedited interpretation reduced mean Ts, Td, and Tb by 99.1%, 99.1%, and 87.2%, respectively.

**CONCLUSION:** Integrating an AI risk model into mammography workflow is feasible and enables same-day evaluation for high-risk women. This approach markedly shortens time to diagnostic imaging and biopsy for timely breast cancer care.

## INTRODUCTION

Timely follow-up after an abnormal screening mammogram is critical for early breast cancer detection. Screening mammograms are typically interpreted offline in batches after the patient has left the facility. If an abnormality is identified, the patient is contacted to return on a separate day for diagnostic imaging, and, if warranted, scheduled for biopsy at a later time. This multistep process introduces cumulative delays that increase patient anxiety, lower satisfaction, and contribute to delays in cancer care (1–3). Women facing transportation challenges, inflexible work schedules, or caregiving responsibilities are disproportionately affected (4). Black, Hispanic, and Asian women, in particular, experience longer delays to diagnostic evaluation and treatment, which exacerbates disparities in breast cancer outcomes (5–11).

Prior work has demonstrated that same-day evaluation of screening abnormalities improves patient experience and adherence to future screening recommendations (12–14). Expedited workflows have also been associated with reduced disparities in breast cancer care (15–17). However, workflow constraints such as staffing limitations, mammography unit/room availability, and unpredictable daily volumes can make immediate screening interpretation and same-day diagnostic evaluation for all patients impractical or unfeasible in most screening programs. In addition, such an approach may not be cost-effective when applied universally (18,19).

Risk-stratified triage for expedited “on-line” screening and diagnostic mammogram interpretation offers a potential solution. Traditional questionnaire-based risk models such as Gail and Tyrer–Cuzick typically provide 10-year or lifetime risk estimates, but do not calculate short-interval (i.e., 1-year) risk, which is most relevant for expedited interpretation (20). In addition, they are time-consuming to collect, depend on patient recall, and are susceptible to language barriers. Artificial intelligence (AI) risk models that assess risk directly from screening mammograms can output risk scores in real-time without additional cumbersome steps. By directing limited resources toward women with high 1-year breast cancer risk scores, programs may maintain operational feasibility while offering expedited assessment for those who are most likely to benefit.

To this end, we conducted a prospective study at an urban safety-net hospital to evaluate the feasibility and performance of offering expedited screening mammogram assessment to women at the top 10th percentile of 1-year breast cancer risk based on an AI risk model. We utilized Mirai, a widely validated mammogram-based AI risk model (21,22). We hypothesized that this strategy would substantially shorten time to screening mammogram result and diagnostic evaluation while preserving workflow capacity by limiting expedited evaluations to a manageable subset of high-risk patients.

## METHODS

### Study Design and Oversight

We conducted a HIPAA-compliant, IRB-approved, prospective, controlled study at an urban safety-net hospital from December 2024 through June 2025. Per the clinicaltrials.gov checklist, this study is exempt from National Clinical Trial (NCT) registration, as it does not satisfy all four required criteria for classification as a clinical trial (23). Informed consent was obtained from all participants.

### AI Risk Algorithm

In the prospective study, the Mirai breast cancer risk model was used to generate 1-year breast cancer risk scores in real-time, immediately after screening mammograms were obtained (21,22). The algorithm processes digital standard craniocaudal (CC) and mediolateral oblique (MLO) views from bilateral 2D screening mammography and outputs non-normalized risk scores. Risk scores were obtained from all patients during the study period. On enrollment days, risk scores were used solely to select patients for enrollment and were not used to guide clinical interpretation or downstream management. On non-enrollment days, radiologists and patients were not notified of flagged studies.

A retrospective analysis of 114,229 local screening mammograms (2006–2023) was performed to calibrate the Mirai risk threshold based on anticipated workflow feasibility. Based on this retrospective cohort, patients in the top 10th percentile of Mirai risk scores had an estimated cancer detection rate (CDR) of approximately 60/1000. Based on an a priori power calculation using cancer detection rates of 60 per 1,000 in the high-risk group and 6 per 1,000 in the standard group, a total sample size of 100 patients for the prospective enrollment was estimated to provide 90% power to detect a significant difference at a two-sided alpha of 0.05.

### Risk Stratification and Recruitment

Eligible participants were women aged 30 years or older who underwent screening mammography and were flagged by the Mirai algorithm as high-risk (top 10th percentile of 1-year risk). Patients were excluded if they had prior history of mastectomy, implants, or were pregnant. On 1–2 designated enrollment days per week, eligible high-risk patients were approached for enrollment. Informed consent was obtained from participants prior to expedited assessments.

### Expedited and Standard Imaging Workflows

On enrollment (intervention) days, consented high-risk women received immediate interpretation of their screening mammogram. Interpreting radiologists were informed that women were prioritized based on Mirai risk, but were not provided the risk scores nor shown any AI localization or heatmaps on the mammograms. At immediate screening mammogram interpretation, patients who received BI-RADS 0 assessments were offered same-day, expedited diagnostic evaluation if feasible based on clinical workflow. If not feasible, diagnostic imaging was scheduled based on next clinical availability. Biopsies were scheduled and performed per standard of care. If warranted, same-day biopsy could be performed at the discretion of the interpreting radiologist and based on staffing availability.

On non-enrollment (control) days, screening examinations followed the standard of care workflow and were read offline after patients left the facility. Women requiring additional imaging were scheduled to return on a separate day for diagnostic evaluation.

On both enrollment and non-enrollment days, standard mammographic views (craniocaudal and mediolateral oblique) were obtained (Selenia Dimensions, Hologic). Images were interpreted by one of 11 board-certified, fellowship-trained breast radiologists with 2-35 years of experience. The BI-RADS assessments at screening mammography and, if performed, diagnostic evaluation were recorded.

### Outcomes and Data Collection, Statistical Analysis

To assess the clinical benefit of AI-based triaging for immediate interpretation, we measured three time intervals: 1) from screening mammogram to screening result (Ts), 2) from screening mammogram to diagnostic evaluation (Td), and 3) from screening mammogram to biopsy (Tb). For expedited patients, the time of screening result release was defined as when results were verbally communicated by the study coordinator to the patient; in the standard workflow for non-enrolled patients, result release was the time the radiologist signed the screening report. For all patients, Td and Tb were defined as the completion times of diagnostic imaging and biopsy examinations, respectively.

Patients were followed for at least three months from the date of their screening mammogram to assess diagnostic and biopsy outcomes. Performance metrics assessed were recall rate, PPV1 (defined as cancers per number of recalled exams), PPV2 (defined as cancers per number of diagnostic exams with a biopsy recommended), PPV3 (defined as cancers per number of biopsies performed), and cancer detection rate (CDR).

To evaluate the clinical feasibility of the AI-based expedited workflow, we recorded how many enrolled patients received immediate interpretation of their screening mammogram. Among patients who received a BI-RADS 0 assessment, we recorded whether diagnostic imaging was completed on the same day. If immediate screening reads or, when recommended, same-day diagnostic evaluation could not be performed, the reason was recorded: clinical workflow constraints (e.g. radiologist or technologist unavailability) or patient-related factor (e.g. patient unavailability).

Medians with interquartile ranges were reported as the primary summary statistics given the skewed distributions of time intervals. Means with 95% confidence intervals were estimated. Group comparisons of Ts, Td, and Tb were performed with the Mann–Whitney U test, and differences in means were evaluated using Welch’s two-sample *t* test. Group comparisons for recall rate, CDR, and PPV1–3 were performed using chi-square tests; Fisher’s exact test was used in comparisons with small counts.

## RESULTS

During the study period, 4145 screening mammograms were performed in 4145 women. Of these, 973 (23.5%) were performed on enrollment days and 3172 (76.5%) were performed on control days. Demographics were similar between enrollment and control days (Table 1).

**Table 1.**
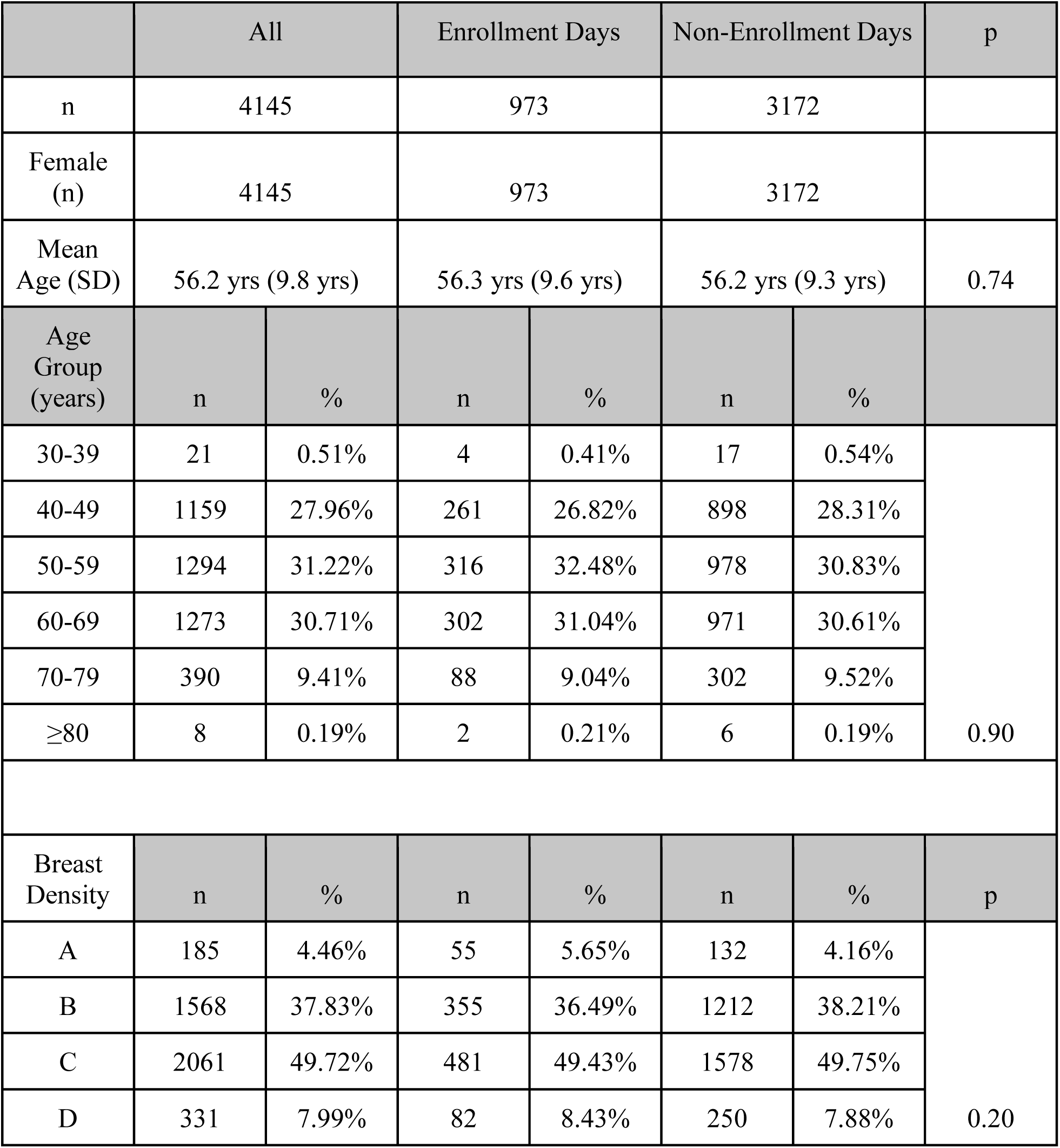
Demographics of Enrollment versus Non-enrollment Days.

|  | All |  | Enrollment Days |  | Non-Enrollment Days |  | p |
| --- | --- | --- | --- | --- | --- | --- | --- |
| n | 4145 |  | 973 |  | 3172 |  |  |
| Female (n) | 4145 |  | 973 |  | 3172 |  |  |
| Mean Age (SD) | 56.2 yrs (9.8 yrs) |  | 56.3 yrs (9.6 yrs) |  | 56.2 yrs (9.3 yrs) |  | 0.74 |
| Age Group (years) | n | % | n | % | n | % |  |
| 30-39 | 21 | 0.51% | 4 | 0.41% | 17 | 0.54% | 0.90 |
| 40-49 | 1159 | 27.96% | 261 | 26.82% | 898 | 28.31% |  |
| 50-59 | 1294 | 31.22% | 316 | 32.48% | 978 | 30.83% |  |
| 60-69 | 1273 | 30.71% | 302 | 31.04% | 971 | 30.61% |  |
| 70-79 | 390 | 9.41% | 88 | 9.04% | 302 | 9.52% |  |
| ≥80 | 8 | 0.19% | 2 | 0.21% | 6 | 0.19% |  |
| Breast Density | n | % | n | % | n | % | p |
| A | 185 | 4.46% | 55 | 5.65% | 132 | 4.16% | 0.20 |
| B | 1568 | 37.83% | 355 | 36.49% | 1212 | 38.21% |  |
| C | 2061 | 49.72% | 481 | 49.43% | 1578 | 49.75% |  |
| D | 331 | 7.99% | 82 | 8.43% | 250 | 7.88% |  |

Using the pre-calibrated threshold, the Mirai model flagged 525 (12.7%) of all 4145 patients as high-risk (i.e. top 10th percentile of 1-year risk). Of the 973 women on enrollment days, 115 (11.8%) were flagged as high-risk. Of the 3172 women on control days, 410 (12.9%) were flagged as high-risk. There was no significant difference in proportion of women flagged as high-risk on enrollment versus non-enrollment days (p=0.41).

Of the 115 patients flagged as high-risk on enrollment days, 2 were not eligible due to participation in another clinical trial. Five could not be reached in time by the research coordinator while they were consenting or coordinating care for another patient. Eight patients did not consent to receive an immediate screening read. In total, 100 patients consented to expedited assessment. Of note, one patient was consented the morning after her screening mammogram owing to a one-time technical problem that delayed transmission of the high-risk notification to the research coordinator until the following day, which was also an enrollment day. She was contacted when the high-risk notification was received, provided informed consent, and returned for same-day expedited diagnostic assessment as her screening mammogram was assessed as BI-RADS 0.

Outcomes in High-Risk Enrolled vs. High-Risk Control Patients

Of the 100 enrolled patients, 94 received immediate screening mammogram interpretation. The 6 patients who did not receive immediate screening interpretation due to workflow constraints had their screening exams interpreted by standard workflow; all 6 were assessed as BI-RADS 1 or 2. Of the 94 patients who received immediate screening mammogram assessments, 31 were assessed as BI-RADS 0. Of those, 30 underwent same-day or next-available scheduled diagnostic evaluation. One patient was scheduled for diagnostic evaluation but did not return. At diagnostic evaluation, 14 were assessed as BI-RADS 1 or 2, 3 were assessed as BI-RADS 3, 12 as BI-RADS 4, and 1 as BI-RADS 5. All 13 women assessed as BI-RADS 4 or 5 underwent biopsy, yielding 6 malignant, 2 high-risk, and 5 benign pathologies (Figure 1). Of the 6 malignant pathologies, 4 were invasive carcinomas (Figure 2) and 2 were ductal carcinomas in situ (Figure 3).

**Figure 1.**
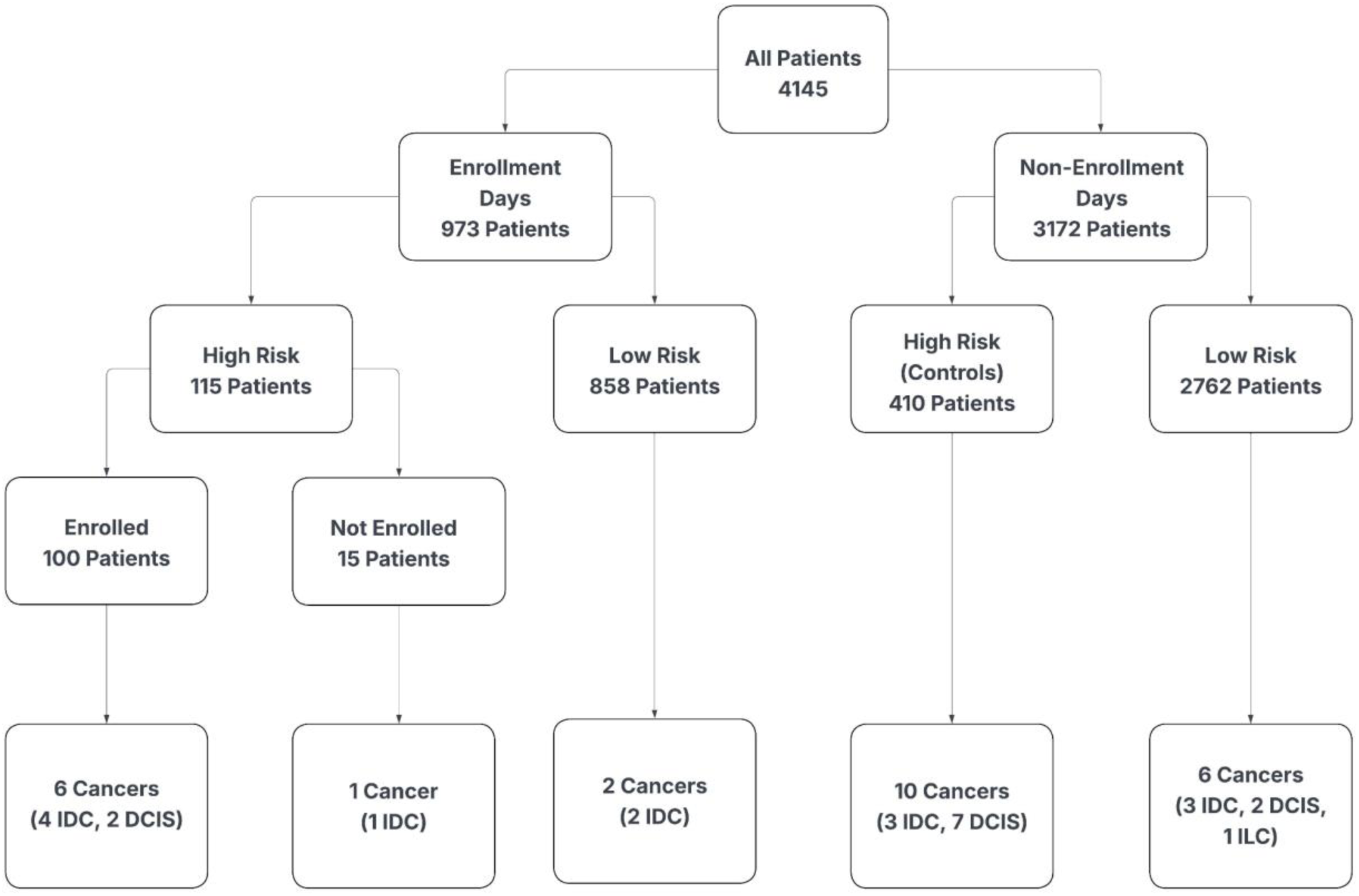
Cancer Outcomes and Mirai Risk Status on Enrollment and Non-enrollment days

**Figure 2A.**
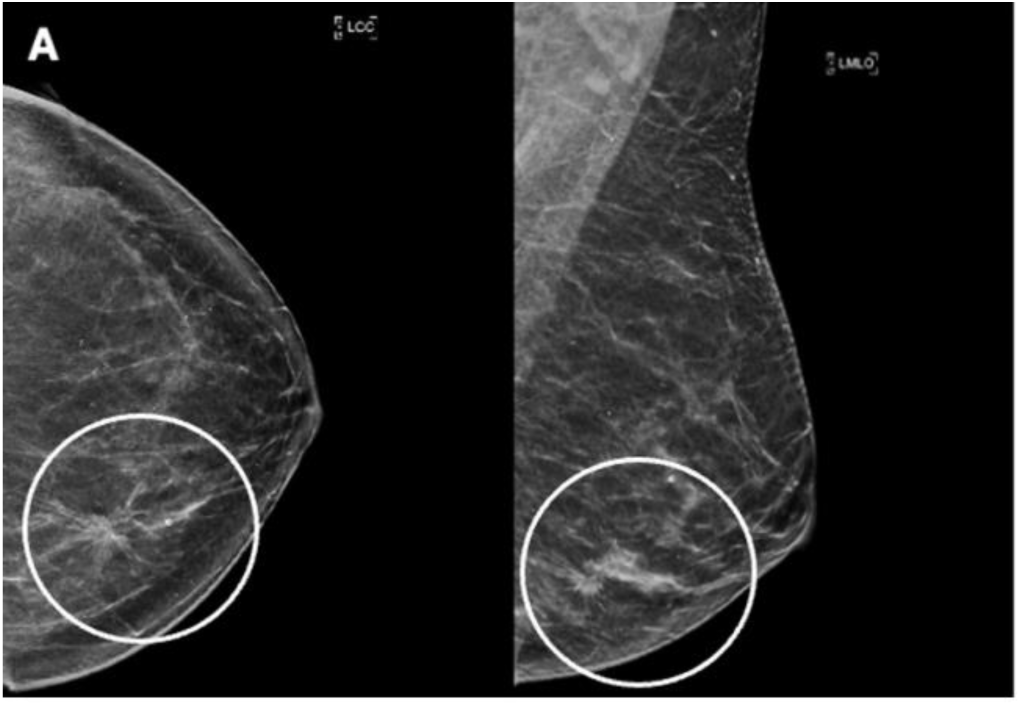
Women in her 40’s presenting for screening mammogram. Left craniocaudal and mediolateral oblique views from screening mammogram flagged as high-risk by Mirai. Immediate screening mammogram assessment identified architectural distortion with associated calcifications in the left lower inner quadrant (circle).

**Figure 2B.**
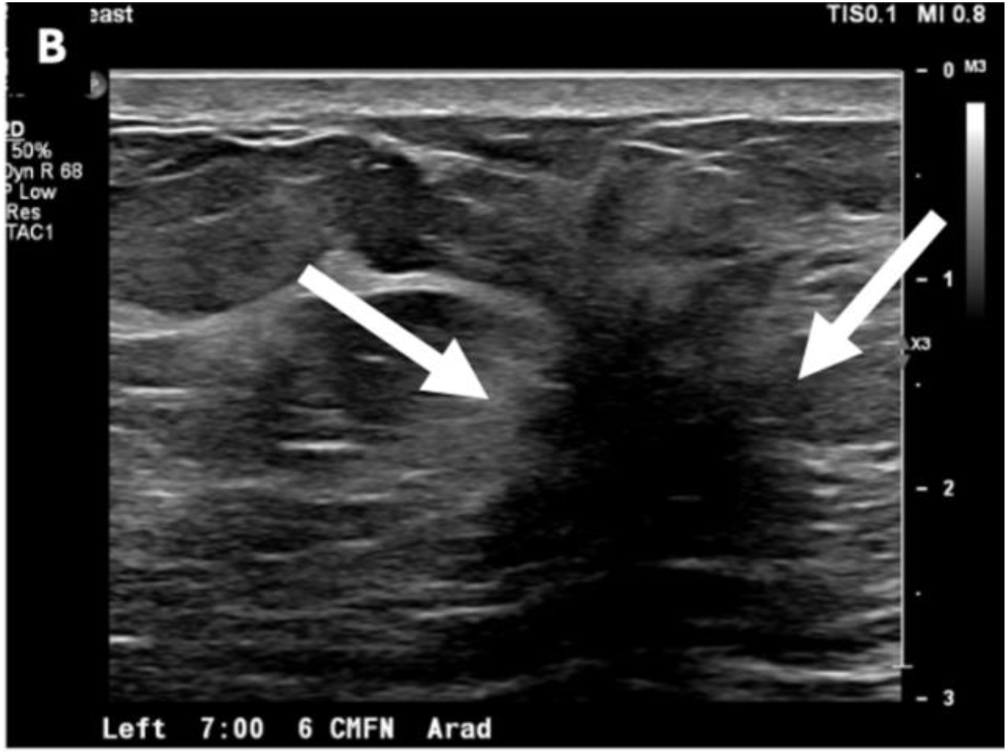
Same day diagnostic mammogram and ultrasound were performed. Diagnostic left breast ultrasound demonstrated architectural distortion spanning 7:00, 4-6 cm from the nipple. Within the architectural distortion, there are two discrete similar appearing masses, the dominant (arrows) measuring up to 1.4 cm, assessed as BI-RADS 4.

**Figure 2C.**
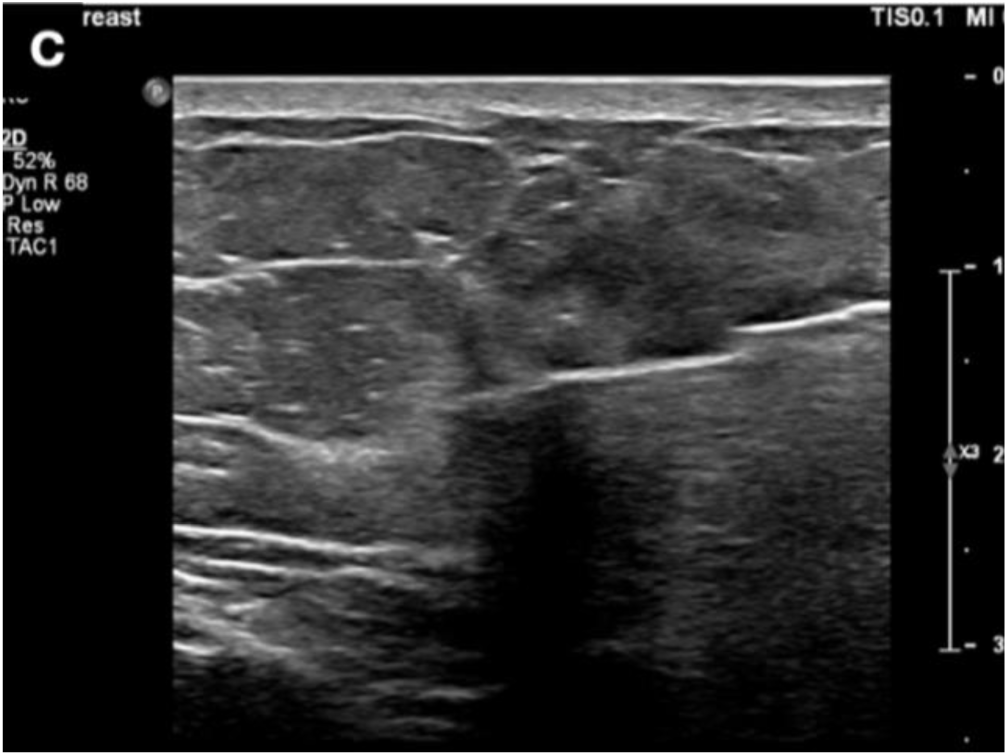
Based on level of suspicion and staffing availability, same day ultrasound guided core biopsy was performed. Pathology yielded invasive ductal carcinoma.

**Figure 2D.**
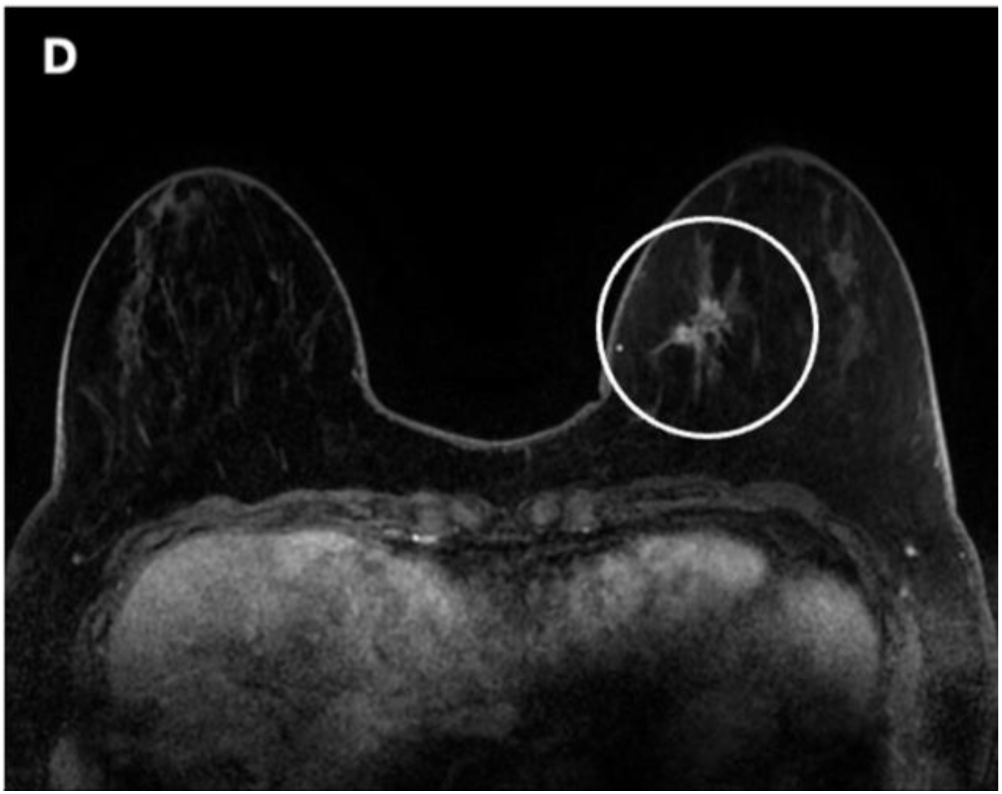
Contrast enhanced breast MRI performed 9 days later demonstrated a 2.6 cm irregularly shaped mass (circle) with spiculated margins and heterogeneous internal enhancement in the left breast, lower inner quadrant. Kinetics curve assessment demonstrated a fast initial and delayed persistent enhancement pattern.

**Figure 3A.**
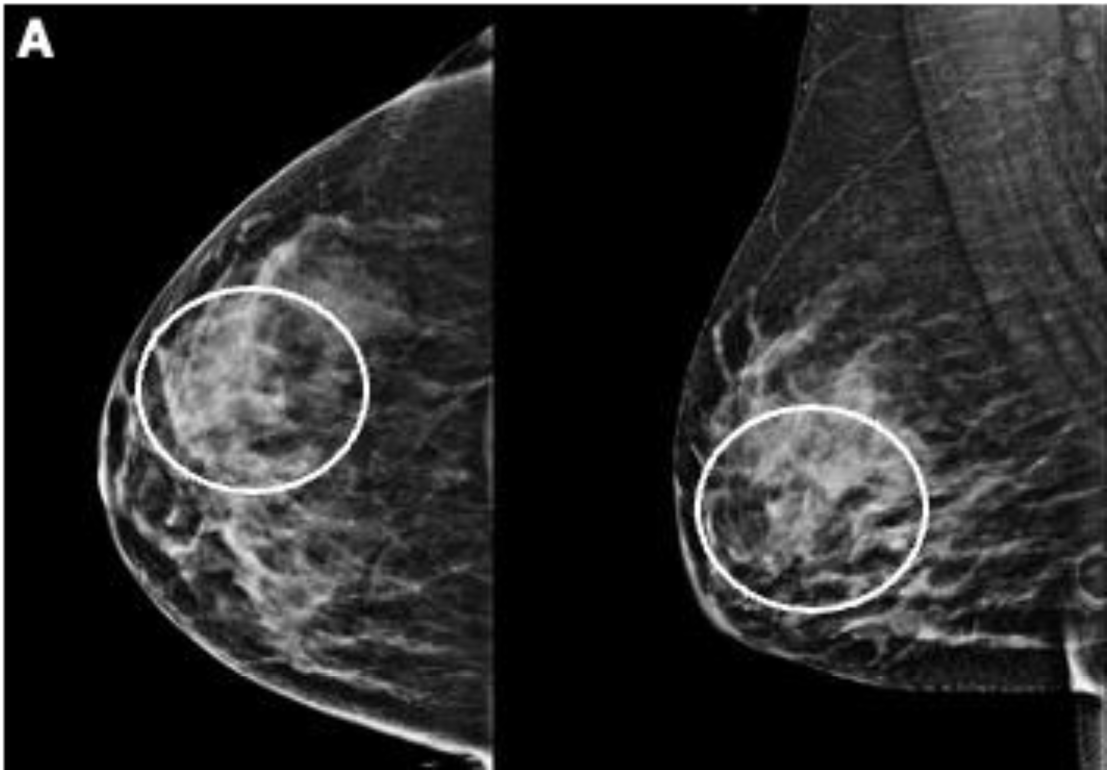
Woman in her 40’s presenting for baseline screening mammogram. Craniocaudal and mediolateral oblique mammograms demonstrate calcifications (circles) in the right outer central breast.

**Figure 3B.**
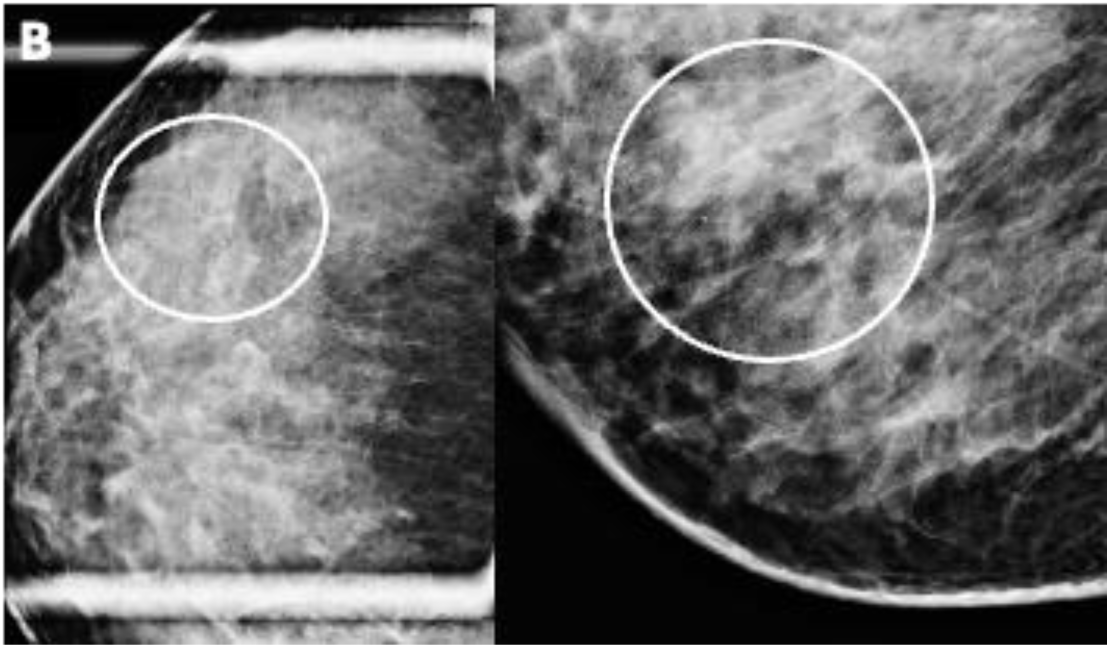
Same day diagnostic evaluation was performed. Spot magnification craniocaudal and lateral views demonstrate amorphous calcifications (circles) in linear distribution in the right breast, outer central quadrant, anterior to middle third, spanning 25 mm, assessed as BI-RADS 4.

**Figure 3C.**
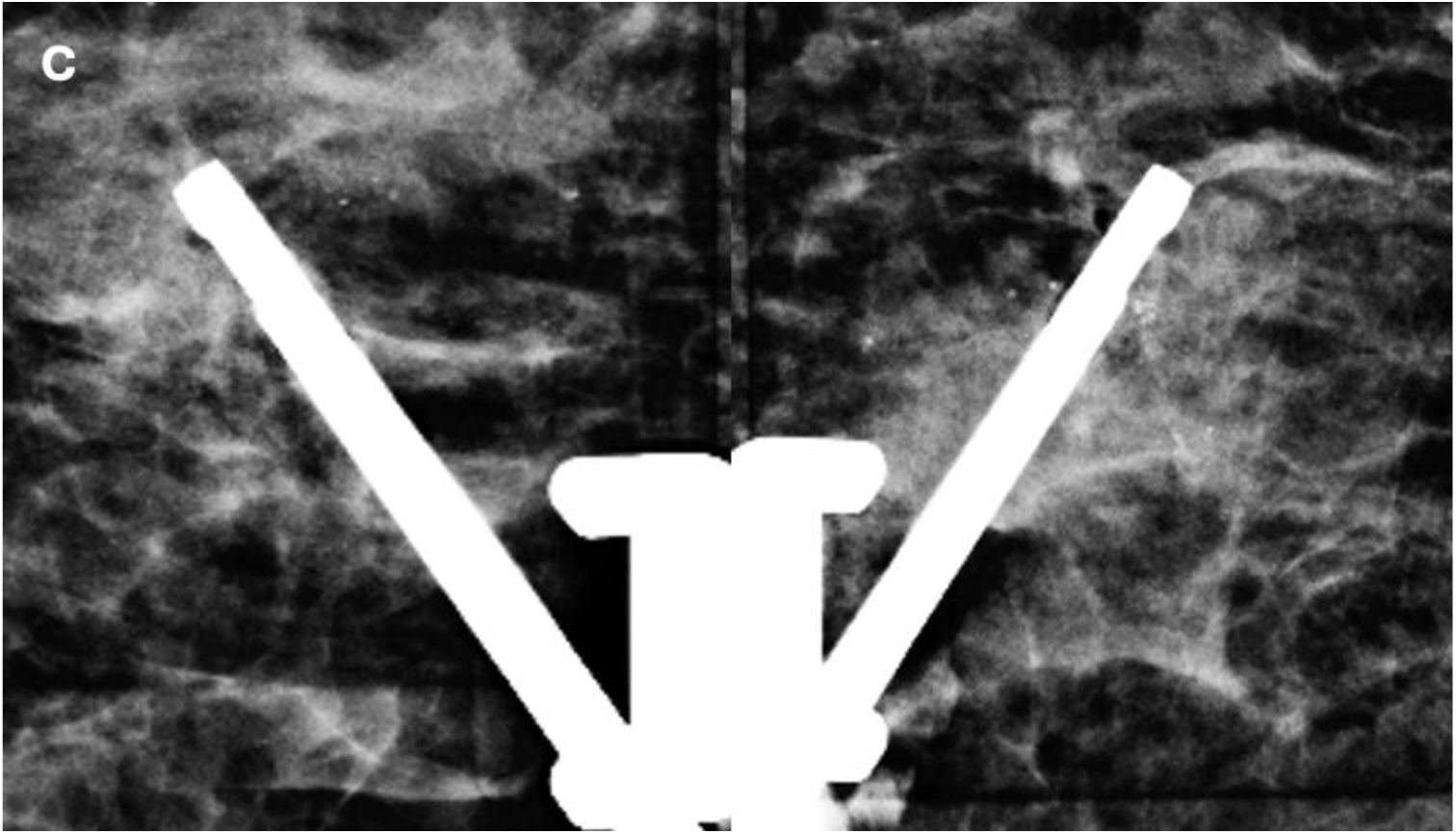
Stereotactic biopsy of the calcifications yielded ductal carcinoma in situ.

**Figure 3D.**
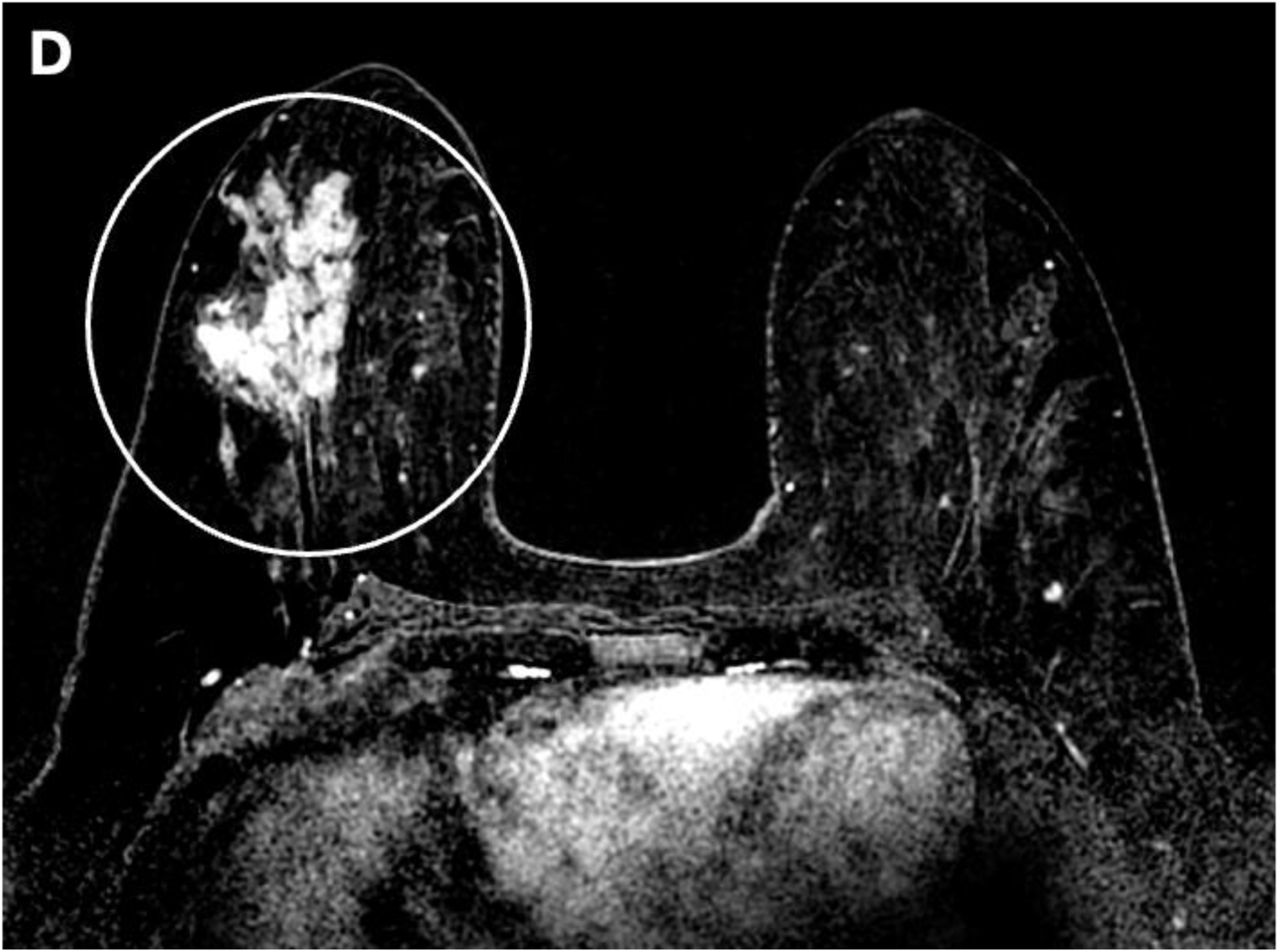
Contrast enhanced breast MRI was performed. Subtraction images demonstrate 10.3 cm non-mass enhancement (circle) with linear distribution and clumped internal enhancement in the right breast, outer central breast. Kinetic curve assessment demonstrates fast initial and delayed washout enhancement pattern

The CDR for this high-risk cohort was 60.0/1000 (6/100) (95% CI 22.3, 126.0), which was significantly higher compared to CDR of 2.3/1000 (2/858)(95% CI 0.28, 8.4)(p<0.001) in the non-high-risk group on enrollment days. Odds ratio was 27.1 (95% CI 4.8, 276.8). PPV1, PPV2, and PPV3 for the high-risk enrolled patients were 19.4% (6/31), 46.2% (6/13), and 46.2% (6/13), respectively (Table 2). Using an intention-to-treat approach, immediate interpretation of every 16.7 AI-flagged screening mammograms resulted in expedited care for a patient with a subsequent screen-detected cancer (6 cancers among 100 flagged high-risk patients). Likewise, offering expedited diagnostic evaluation to every 5.2 recalled high-risk patients resulted in one additional cancer being diagnosed more promptly (6 cancers among 31 recalled, high-risk patients).

**Table 2.** Screening Performance Metrics in Consented High-risk vs. High-risk Controls.

|  | All | Expedited High-Risk | High-Risk Controls | p |
| --- | --- | --- | --- | --- |
| n | 4145 | 100 | 410 |  |
| Recall Rate | 12.8%<br>(532/4145) | 31.0% (31/100) | 21.5% (88/410) | 0.04* |
| CDR | 6.0/1000<br>(25/4145) | 60.0/1000 (6/100) | 24.4/1000 (10/410) | 0.10 |
| %<br>Diagnostic<br>Exams<br>Performed | 89.1%<br>(474/532) | 96.8% (30/31) | 89.8% (79/88) | 0.45 |
| Diagnostic<br>Assessment |  |  |  |  |
| BI-RADS<br>1 or 2 | 47.7%<br>(226/474) | 46.7% (14/30) | 31.7% (25/79) | 0.14 |
| BI-RADS<br>3 | 22.4%<br>(106/474) | 10.0% (3/30) | 19.0% (15/79) | 0.26 |
| BI-RADS<br>4 | 29.5%<br>(140/474) | 40.0% (12/30) | 48.1% (38/79) | 0.45 |
| BI-RADS<br>5 | 0.4% (2/474) | 3.3% (1/30) | 1.3% (1/79) | 0.47 |
| PPV1 | 4.7%<br>(25/532) | 19.4% (6/31) | 11.4% (10/88) | 0.26 |
| PPV2 | 17.6%<br>(25/142) | 46.2% (6/13) | 25.6% (10/39) | 0.17 |
| PPV3 | 18.3%<br>(25/137) | 46.2% (6/13) | 27.0% (10/37) | 0.20 |

Of the 410 high-risk control patients on non-enrollment days, 21.5% were recalled (88/410), which was significantly lower compared to that in high-risk enrolled patients (31/100; p=0.04). Of those, 79 underwent diagnostic evaluation. Fifteen were assessed as BI-RADS 3, 38 as BI-RADS 4, and 1 as BI-RADS 5. The CDR for this cohort was 24.4/1000 (10/410; 95% CI 11.8, 44.4), which did not differ significantly from that of the high-risk enrolled group (p=0.10).

PPV1, PPV2, and PPV3 for high-risk controls were 11.4% (10/88), 25.6% (10/39), and 27.0% (10/37), respectively (Table 2).

Across both enrollment and non-enrollment days, the Mirai model flagged 68.0% (17/25) of screen-detected cancers. Of the 15 patients who were flagged as high-risk on enrollment days but who were not enrolled, one resulted in cancer (invasive ductal carcinoma). On enrollment days, Mirai flagged 77.8% (7/9) of screen-detected cancers. On non-enrollment days, it flagged 62.5% (10/16) of screen-detected cancers.

### Time to Screening Result, Diagnosis, and Biopsy

Median time from screening mammogram to screening result (Ts) was significantly shorter for all enrolled expedited patients (13.0 minutes vs. 191.9 minutes; p<0.001) and for enrolled expedited patients with screen-detected cancers (12.23 minutes vs. 655.86 minutes; p<0.001) compared to high-risk controls on non-enrollment days (Tables 3 and 4). Median time from screening mammogram to completion of diagnostic evaluation (Td) was significantly shorter for all enrolled expedited patients (1.27 hours vs. 852.78 hours; p<0.001) and for expedited patients with screen-detected cancers (1.25 vs. 1535.85 hrs; p=0.003) compared to high-risk controls.

**Table 3.** Workup Times for Expedited High-Risk Patients vs. High-Risk Controls.

|  |  | Expedited<br>High-Risk<br>Patients | High-Risk<br>Controls | p-value |
| --- | --- | --- | --- | --- |
| Screening Mammogram to<br>Screening Result (minutes) | n | 94 | 410 |  |
|  | **Median** | **13.0** | **191.9** | **<0.001** |
|  | IQR | 9.2, 19.8 | 84.1, 1207.8 |  |
|  | **Mean** | **14.7** | **900.9 (15.0**<br>**hours)** | **<0.001** |
|  | 95% CI | 13.2, 16.1 | 769.8, 1030.8 |  |
| Screening Mammogram to<br>Completion of Diagnostic<br>Evaluation (hours) | n | 30 | 79 |  |
|  | **Median** | **1.27** | **852.8 (35**<br>**days 12.78**<br>**hours)** | **<0.001** |
|  | IQR | 0.9, 2.2 | 498.3, 1661.0 |  |
|  | **Mean** | **16.0** | **1078.4 (44**<br>**days 22.4**<br>**hours)** | **<0.001** |
|  | 95% CI | -4.3, 36.3 | 872.9, 1283.9 |  |
| Screening Mammogram to<br>Completion of Biopsy<br>(days) | n | 13 | 37 |  |
|  | **Median** | **20.1** | **59.0** | **<0.001** |
|  | IQR | 5.2, 28.1 | 38.0, 96.7 |  |
|  | **Mean** | **18.4** | **69.9** | **<0.001** |
|  | 95% CI | 9.2, 27.6 | 54.8, 85.0 |  |

**Table 4.**
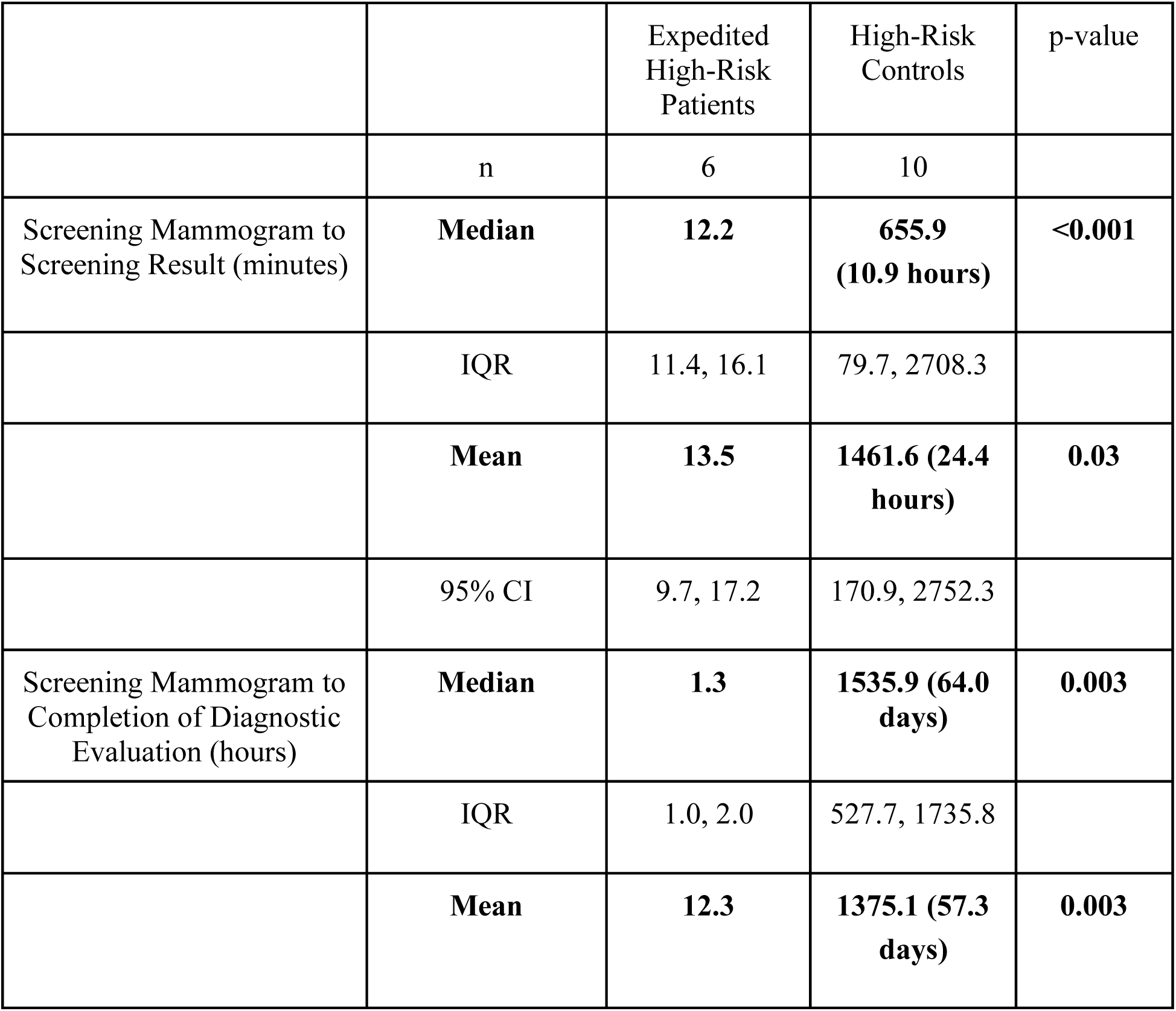
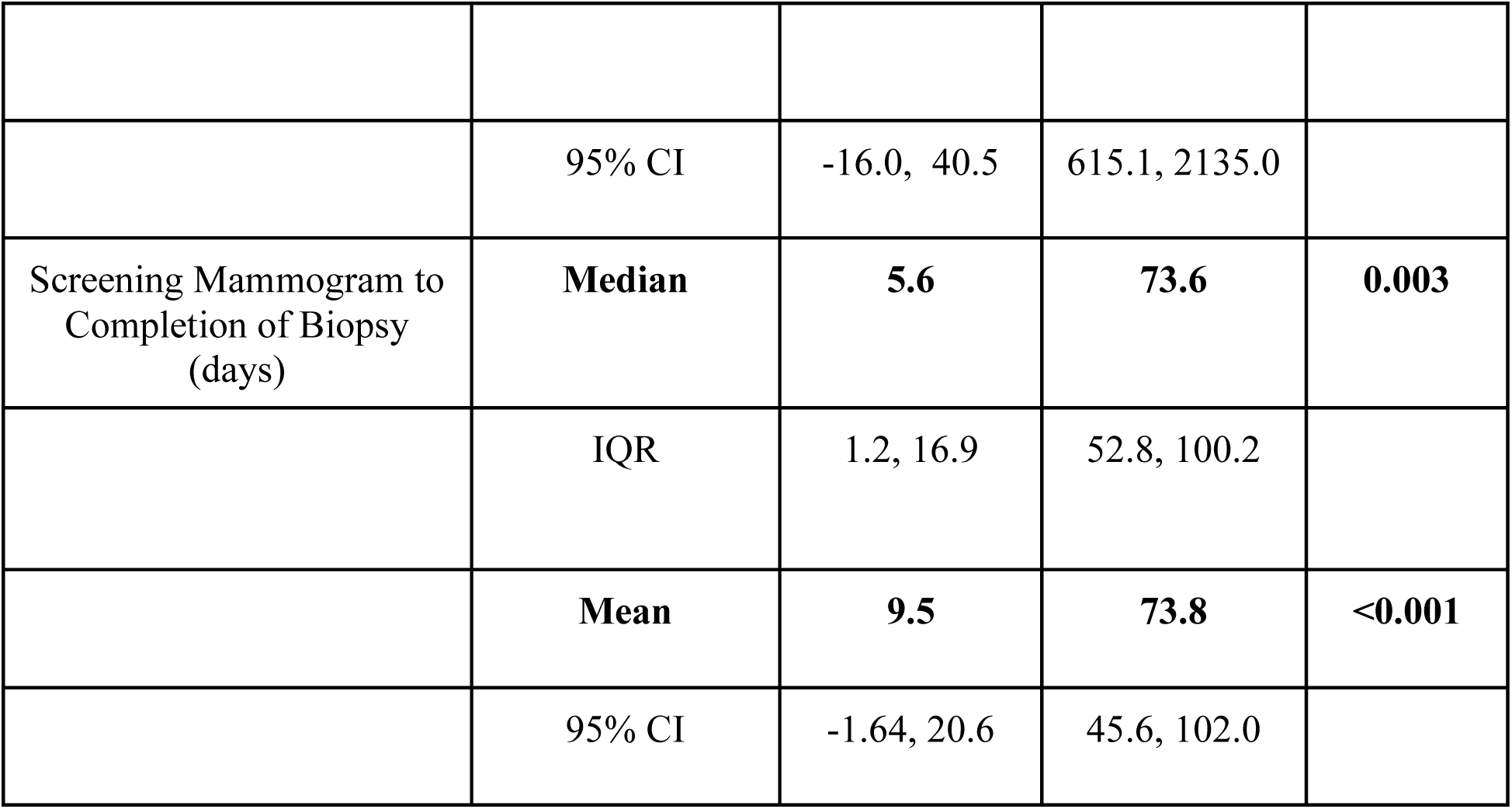
Workup Times for Screen-Detected Cancers.

Median time from screening mammogram to completion of biopsy was significantly shorter for all expedited patients (20.11 days vs. 58.96 days; p<0.001) and for expedited patients with screen-detected cancers (5.61 vs. 73.60 days).

Similarly, mean times were significantly shorter for expedited patients compared to high-risk controls on non-enrollment days (Tables 3 and 4)(Figures 4, 5, and 6). For all expedited patients, mean Ts was reduced by 98.3% (14.7 vs 900.9 minutes; p<0.001), Td by 98.5% (16.0 vs 1078.4 hours; p<0.001), and Tb by 73.7% (18.4 vs 69.9 days; p<0.001). Among expedited patients with screen-detected cancers, reductions were even greater: Ts was reduced by 99.1% (13.5 vs 1461.6 minutes; p=0.03), Td by 99.1% (12.3 vs 1375.1 hours; p=0.003), and Tb by 87.2% (9.5 vs 73.8 days; p<0.001).

**Figure 4.**
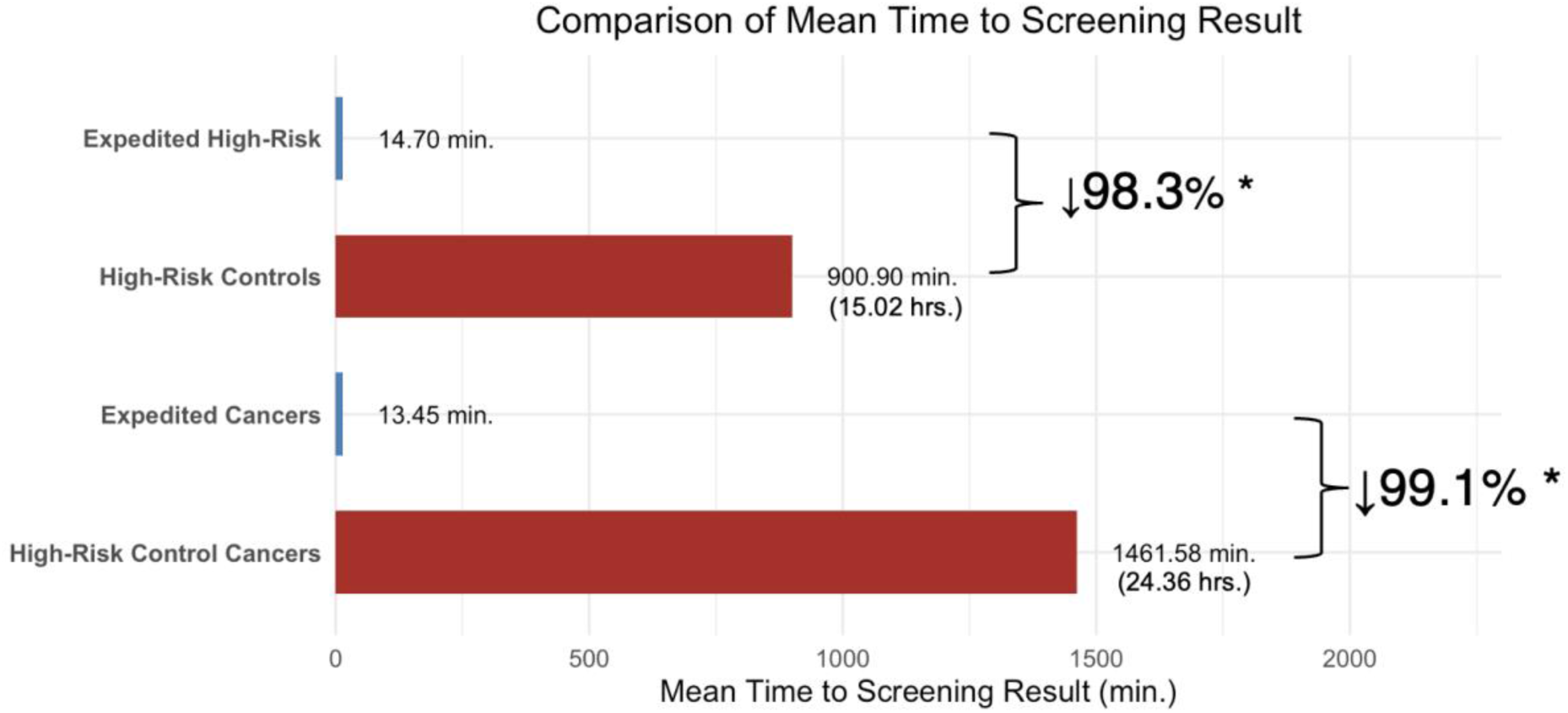
Comparison of Mean Time to Screening Result (Ts)

**Figure 5.**
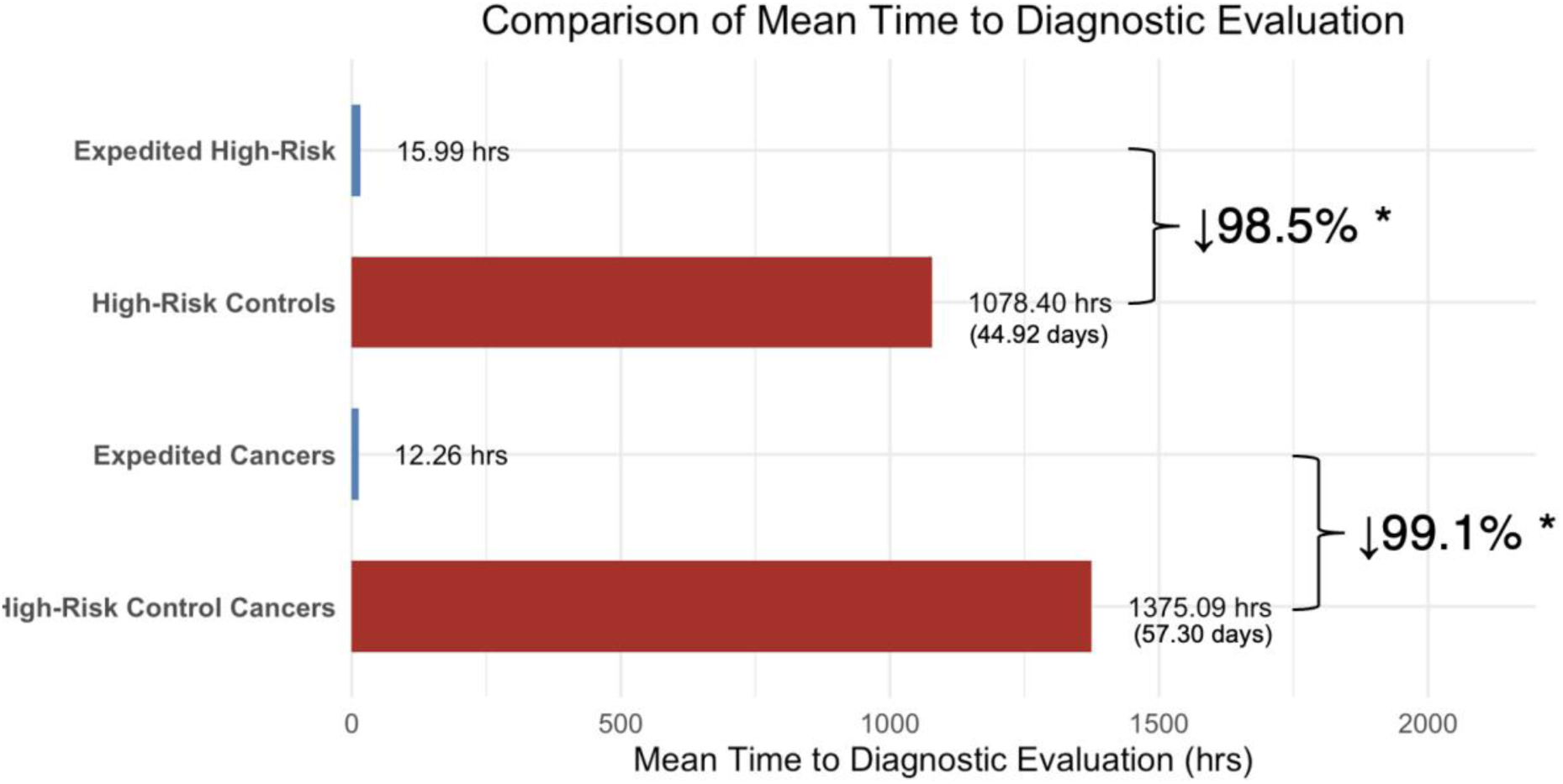
Comparison of Mean Time to Diagnostic Evaluation (Td)

**Figure 6.**
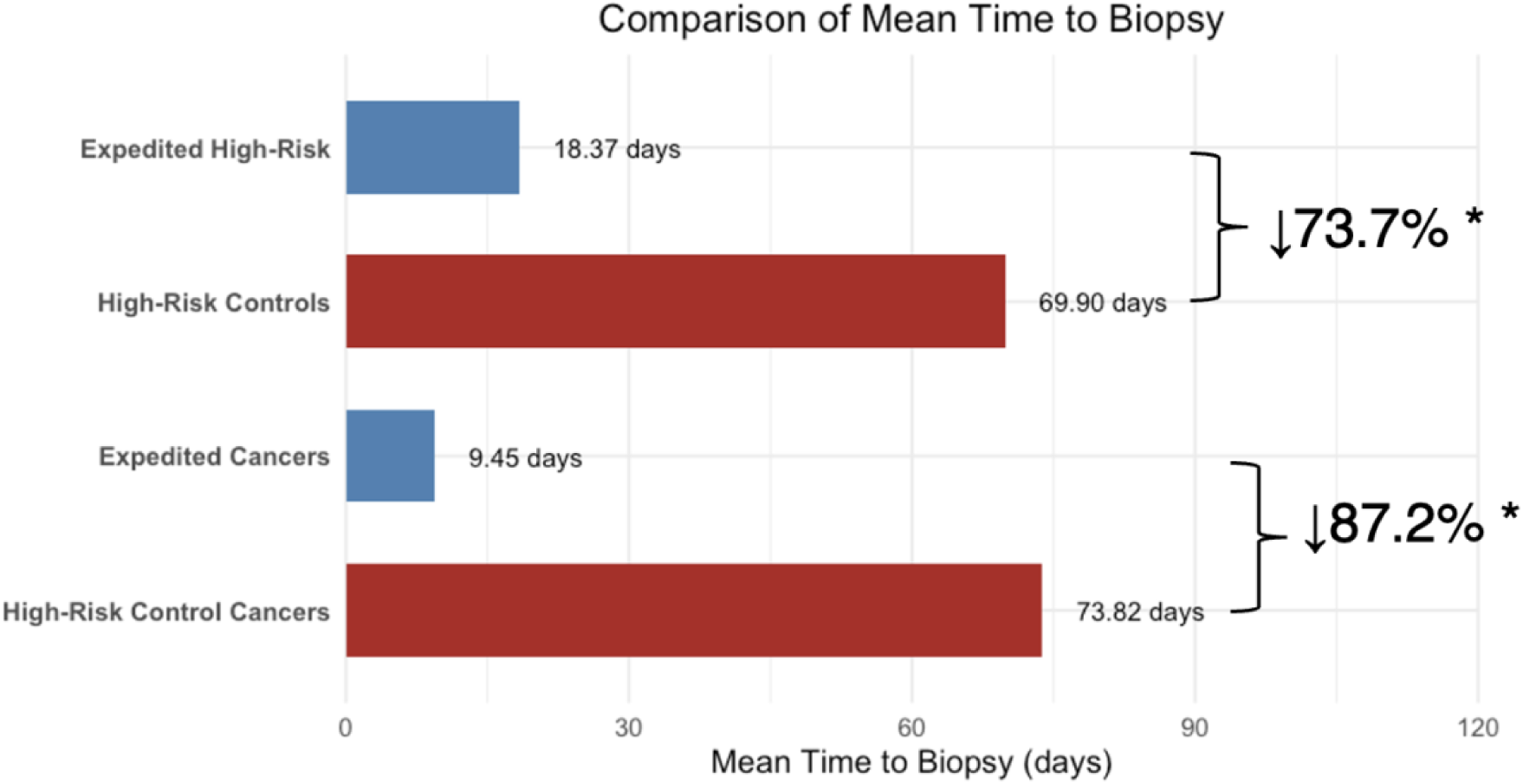
Comparison of Mean Time to Biopsy (Tb)

### Workflow Feasibility

Of the 100 enrolled patients, 94 received immediate screening interpretation; six did not receive immediate reads due to clinical time constraints (n=4) and wait time (n=2). Among patients who received immediate screening interpretation, 97.9% (92/94) received their screening result within 30 minutes. Of the 31 enrolled patients who were assigned BI-RADS 0 assessment at immediate screening interpretation, 26 (83.9%) underwent diagnostic imaging on the same day. Five (16.1%) required scheduling on a different day due to workflow limitations. Of these, 4 received their diagnostic evaluation 1, 2, 3, and 12 days later, respectively. One did not return for diagnostic evaluation. Two enrolled patients who underwent same-day diagnostic imaging also underwent same-day biopsy at the discretion of the interpreting radiologist.

## DISCUSSION

In this prospective study, integration of an AI-based risk model for triaging enabled immediate interpretation and expedited diagnostic evaluation for women with high 1-year risk scores. The most pronounced benefit was observed among expedited patients with screen-detected cancers, for whom time to screening result, diagnostic result, and biopsy were reduced by 99.1%, 99.1%, and 87.2%, respectively, compared to control patients on non-enrollment days. Such reductions have the potential to accelerate treatment and improve clinical outcomes (24). For high-risk patients with negative exams, expedited interpretation can shorten the period of uncertainty, improve the screening experience, and potentially improve adherence to annual screening recommendations in high-risk women (13).

Our results highlight the importance of thoughtful, workflow-oriented design to enable feasible implementation. Threshold selection is central to the performance of risk-stratification strategies: flagging too large a proportion of patients risks overwhelming diagnostic capacity, whereas flagging too few may limit benefit. In our study, applying a 10th-percentile Mirai threshold for 1-year risk identified 77.8% of screen-detected cancers while flagging 12.7% of patients for expedited evaluation. Nearly all flagged patients (94%) were able to receive immediate screening reads and most (83.8%) completed same-day diagnostic evaluation, supporting the operational feasibility of this approach.

These findings build on prior work by Friedewald et al., which used a commercial AI to triage mammograms for expedited evaluation (25). While they retrospectively calibrated their threshold to identify 14% of patients as high-risk, the threshold flagged 29% of patients in prospective implementation. We prospectively applied the 10th-percentile threshold determined from retrospective validation, and the observed rate of 12.7% aligned closely with the anticipated proportion. This more specific threshold promotes workflow feasibility. In our study, 83.9% (26/31) of patients with BI-RADS 0 findings at immediate screening mammography received same-day diagnostic evaluation, compared with 72.7% (24/33) reported by Friedewald et al. In our study, immediate interpretation of every 16.7 AI-flagged mammograms (6 cancers among 100 flagged) resulted in expedited care for one patient with screen-detected cancer, compared with 43.7 mammograms (3/131) in their study. Similarly, offering expedited diagnostic evaluation to every 5.2 recalled high-risk patients (6/31) yielded one additional promptly diagnosed cancer, versus 11.0 (3/33) in their study. This supports that our 10th percentile threshold using the Mirai model is a more efficient strategy to prioritize patients with cancer.

Notably, recall rates among high-risk enrolled patients in our study was high (31.0%). This likely reflects the higher proportion of cancers in this patient subset as well as the prospective design. Radiologists may have been more inclined to recommend additional imaging when they are aware of the patient’s elevated risk status and when the patient was already present, eliminating the need for a return visit. Importantly, despite this high recall rate, PPV1–3 remained high (19.4%, 46.2%, and 46.2%, respectively) compared with that in the overall population (4.70%, 17.61%, and 18.25%) and high-risk controls (11.36%, 25.64%, and 27.03%), supporting that the elevated recall rate reflected appropriate identification of cancers in high-risk patients rather than unnecessary workup.

By prospectively validating AI-based triaging for screening programs, this study provides evidence for how risk-based tools can be translated into clinical workflows. Our findings are particularly relevant at a time when many commercial AI applications are increasingly marketed directly to patients on a pay-for-access basis, raising concerns that such approaches may exacerbate socioeconomic inequities in breast cancer care. In contrast, we demonstrate the integration of Mirai, a free and open-source AI model, within existing clinical workflows. By lowering barriers to patient access, this approach may help reduce disparities and narrow gaps in breast cancer outcomes. The clinical implications of these findings may be greatest in safety-net settings, where vulnerable populations often face barriers to timely follow-up and are at higher risk of being lost to care. By helping patients who are most likely to have cancer receive expedited evaluation, AI-based triage may mitigate disparities in access to and timeliness of care and improve outcomes (15,17,24).

Our study has several limitations. First, it was conducted at a single safety-net institution, potentially limiting generalizability. Second, workflow feasibility is likely to vary across sites and will depend on each breast screening program’s volumes and operational capacity. Third, patients were followed for at least three months to capture diagnostic and biopsy outcomes; cancers diagnosed beyond this interval were not included. Finally, patients with a history of mastectomy, current pregnancy, or breast implants were not enrolled.

In summary, this prospective study demonstrates that an AI-based risk model can be feasibly integrated into screening workflows to enable immediate interpretation and same-day diagnostic evaluation for high-risk women. This approach substantially reduced delays to diagnosis and biopsy, with potential to improve outcomes, lessen anxiety, and support adherence to follow-up. By prospectively validating both predictive performance and workflow feasibility, our findings provide a framework for translating AI risk-stratification tools into screening programs.

## Data Availability

All data produced in the present study are available upon reasonable request to the authors.

